# Uterine Fibroids Among Caribbean and African Women in the UK: A Rapid Scoping Review

**DOI:** 10.1101/2025.04.15.25325785

**Authors:** F Ruddock, D Anumba, AL Kwansa, P Walters, M Ejim, L Adjei-Mensah

## Abstract

**Background:** Uterine fibroids (leiomyomas) are common benign tumours affecting women in the UK, with women of African and Caribbean descent reporting a higher prevalence, early onset, and greater symptom severity compared to other ethnic groups. Despite this known disparity, there is a paucity of specific evidence regarding the prevalence, risk factors, management, and outcomes for this population within the UK healthcare context.

**Objective:** To conduct a rapid scoping review aimed at identifying, mapping, and synthesising the existing UK-based evidence concerning uterine fibroids among women of African and/or Caribbean ancestry.

**Methods:** A search strategy was developed and used in MEDLINE, CINAHL, and the Cochrane Library to identify empirical studies conducted in the UK and reporting on any aspect of uterine fibroids (genetics, risk factors, clinical presentation, treatment, quality of life (QoL), complications, healthcare access, patient experiences, etc) in self-identified African and/or Caribbean women. Grey literature sources were also included in the searches. Data from the included articles and reports were extracted and synthesized narratively.

**Results:** The search identified six studies conducted between 2014 and 2021. These included 1 genome-wide association study (GWAS), 1 clinical trial, 2 observational studies, and 2 case reports. Key findings suggest a genetic predisposition to uterine fibroids, with GWAS data identifying shared and ancestry-specific loci relevant to UK women of African ancestry. Treatment evidence also showed that compared to Uterine Artery Embolisation (UAE), myomectomy offered better QoL outcomes, especially for women seeking uterine preservation. There was also evidence showing longer post-surgical recovery times for Black women after myomectomy, as well as reports of rare but severe complications associated with the condition. The potential role of Vitamin D deficiency as a risk factor for uterine fibroids among Black women in the UK was also reported.

**Conclusion:** This review confirms the significant burden of uterine fibroids among African and Caribbean women in the UK and highlights a critical lack of focused research. Research priorities should include large-scale epidemiological studies to understand the risk or protective factors associated with the progression of the condition (including the role of Vitamin D). Future work should also include comparative effectiveness trials to identify the most relevant treatment options, as well as a further exploration of patient experiences. Our findings underscore the need for culturally competent clinical care, improved patient awareness, equitable access to diverse treatment options, and targeted policy initiatives to address health disparities in uterine fibroid management for Black women in the UK.

## Introduction

Uterine fibroids, also known as leiomyomas or myomas, are benign monoclonal tumours arising from the smooth muscle cells of the uterus (Holdsworth-Carson et al., 2014). They represent the most common gynaecological tumours in women of reproductive age, and although they are non-cancerous, they can cause significant morbidity and negatively impact quality of life (Stewart et al., 2017; Wise & Laughlin-Tommaso, 2016).

The prevalence of uterine fibroids varies significantly based on the population studied, diagnostic methods employed, and age range considered (Stewart et al., 2017; Martín-Merino et al., 2016). Studies utilizing ultrasound screening indicate a remarkably high cumulative incidence, with some reports suggesting that up to 70% of white women and over 80% of Black women will develop fibroids by age 50 (Baird et al., 2003). Self-reported data, although potentially underestimating the true prevalence, also show substantial rates, with age-specific incidence peaking during the premenopausal years (Wise et al., 2005; Marshall et al., 1997).

A complex interplay of factors contributes to the development of uterine fibroids. Age is a crucial determinant, with incidence rates increasing from menarche to menopause (Wise & Laughlin-Tommaso, 2016). Race and ethnicity are also major factors, with Black women experiencing significantly higher incidence, earlier onset, and greater severity of fibroids compared to women of other racial groups (Baird et al., 2003; Marshall et al., 1997; Wise et al., 2005). Reproductive factors also play a role; parity appears to be protective, although the precise mechanisms underlying this association remain unclear (Baird & Dunson, 2003).

Lifestyle and environmental factors have also been implicated in the development of fibroids. Obesity and increased body fat distribution are consistently associated with an elevated risk (Qin et al., 2021; Lee et al., 2018; Sato et al., 1998). Dietary factors, such as a high consumption of red meat and a low intake of fruits and vegetables, have been suggested to increase risk, while the role of dietary fat remains debated (Wise et al., 2014; He et al., 2013; Wise et al., 2016). Vitamin D deficiency has emerged as a potential risk factor, with numerous studies suggesting an inverse relationship between vitamin D levels and the risk of fibroids (Brakta et al., 2015; Baird et al., 2013; Ciebiera et al., 2018). Early-life exposures, including factors experienced in utero or during childhood, may also influence the development of fibroids later in life (D’Aloisio et al., 2012).

Growing evidence suggests a strong genetic component in fibroid susceptibility. Familial studies have shown a higher incidence of fibroids among relatives, indicating a heritable predisposition (Vikhlyaeva et al., 1995; Luoto et al., 2000). Genome-wide association studies (GWAS) have identified multiple genetic loci linked to fibroid risk, involving genes related to hormone metabolism, cell growth, and the extracellular matrix (Cha et al., 2011; Edwards et al., 2013; Eggert et al., 2012; Hellwege et al., 2017). Specific polymorphisms in genes such as FASN, BET1L, TNRC6B, CYP1B1, and those associated with age at menarche have been correlated with fibroid risk or size (Wise et al., 2016; Gallagher & Morton, 2016; Bideau & Alleyne, 2016; Ponomarenko et al., 2020; Aissani et al., 2015; Mu et al., 2015).

Clinically, fibroids can present a wide range of symptoms, or they may be completely asymptomatic (Foth et al., 2017). Common symptoms include heavy menstrual bleeding (menorrhagia), prolonged menstrual periods, and pelvic pain (Agboola et al., 2021). In some instances, fibroids may contribute to infertility or complications during pregnancy. Although rare, there exists a slight risk of malignant transformation to leiomyosarcoma, underscoring the importance of precise diagnosis and monitoring (Yorgancı et al., 2020).

Popular treatment options for uterine fibroids include medical and surgical interventions. Medical management encompasses hormonal therapies such as GnRH agonists and antagonists, selective progesterone receptor modulators (SPRMs), and levonorgestrel-releasing intrauterine systems (LNG-IUS), along with non-hormonal options like tranexamic acid for managing heavy bleeding. Surgical methods include myomectomy (fibroid removal), hysterectomy (uterus removal), and uterine artery embolization (UAE), as well as newer, less invasive techniques like magnetic resonance-guided focused ultrasound surgery (MRgFUS) and radiofrequency ablation (RFA).

The impact of uterine fibroids extends beyond the individual, imposing a significant economic burden on healthcare systems. Direct costs encompass medical consultations, diagnostic imaging, medications, and surgical procedures (Soliman et al., 2015; Cardozo et al., 2012). Indirect costs, such as lost productivity due to absenteeism and decreased work performance, further add to the overall economic impact (Soliman et al., 2015).

In the UK, although uterine fibroids disproportionately affect Black women, there is a significant paucity of evidence on the prevalence, risk factors, management, and outcomes for this specific population. Treatment practices also vary considerably (Martín-Merino et al., 2016). Addressing this knowledge gap through this rapid review is crucial for informing culturally tailored healthcare strategies, improving clinical practice, and reducing health disparities experienced by Black women in the UK.

## Methods

This rapid scoping review was guided by the methodological framework proposed by Arksey and O’Malley (2005) and further refined by Levac et al. (2010). It was also informed by the Preferred Reporting Items for Systematic Reviews and Meta-Analyses extension for Scoping Reviews (PRISMA-ScR) and recommendations from the Joanna Briggs Institute (JBI) (Peters et al., 2020). Briefly, the frameworks by Arksey and O’Malley (2005), Levac et al. (2010), PRISMA-ScR, and the Joanna Briggs Institute recommend a systematic and transparent approach to reviewing primary data from existing literature, starting with broad research questions, the inclusion of a wide range of sources, and the development of clear protocols that clearly report on the study rationale, objectives, and review findings.

### Literature Search

The study’s research question and recommendations from the JBI informed the choice of the PCC (Population, Concept, Context) search framework, aligning with the scoping nature of this review. An initial internet search was conducted to assess the extent of the available literature and to identify synonyms related to the population (Caribbean and African populations, e.g., “Black”, “African descent”, “Caribbean descent”, “Afro-Caribbean”), Concept (Uterine fibroids, e.g., “leiomyoma”, “myoma”), and Context (United Kingdom, e.g., “UK”, “Great Britain”, “England”, “Scotland”, “Wales”, “Northern Ireland”). This initial search informed the development and refinement of a comprehensive search strategy for use in three relevant electronic databases: MEDLINE (via PubMed), CINAHL, and the Cochrane Library. The final search strategy was adapted as needed for each database. Search terms in the strategy were combined using Boolean operators for literature published in English (Supplementary Material 1). There were no time limits for the searches (i.e., up to February 2025, when they were conducted). Time limits were not imposed because the initial scoping search yielded few relevant results, suggesting that accurately capturing pertinent literature would require extending, rather than limiting, the time scope of the searches.

Grey literature sources were also searched for relevant literature. These included Google Scholar, the websites of the UK’s National Health Service (NHS), the National Institute for Health and Care Excellence (NICE), the National Institute for Health Research (NIHR), the Royal College of Nursing, and the ProQuest theses and dissertation repository.

Finally, the reference lists of all included studies were manually searched to identify any additional relevant articles that may have been missed during the database and grey literature searches. Search outputs were subsequently transported into Covidence for deduplication and screening.

### Eligibility and Study Selection

#### Inclusion/Exclusion Criteria

Studies were included if they were peer-reviewed, empirical reports on women who self-identified or were identified as Afro-Caribbean, i.e., of African and/or of Caribbean descent living in the UK. Studies that reported mixed ethnic populations were included only if data for Caribbean and African women in the UK were separately reported. Studies were also included in the review if they explicitly reported on any aspect of uterine fibroids, including prevalence and incidence, risk factors (genetic, environmental, lifestyle), clinical presentation and symptoms, diagnosis and screening, treatment and management, complications, impact on quality of life, fertility, and pregnancy, psychosocial impact, healthcare utilisation and access, and patient experiences and perspectives. Only studies conducted in the UK or reporting on data collected from Black women living in the UK were included in this review.

Studies were not included if they specifically focused on or reported conditions other than uterine fibroids for Caribbean and African women. Studies were also excluded if they reported on women living outside the UK.

### Screening

Two reviewers independently screened the titles and abstracts of all studies retrieved after literature searches had been completed. Potentially relevant studies were subjected to full-text review by two independent reviewers. Disagreements were resolved through discussions or consultations with a third reviewer. The study selection process has been documented in the PRISMA flow diagram in Figure 1.

**Figure 1.**
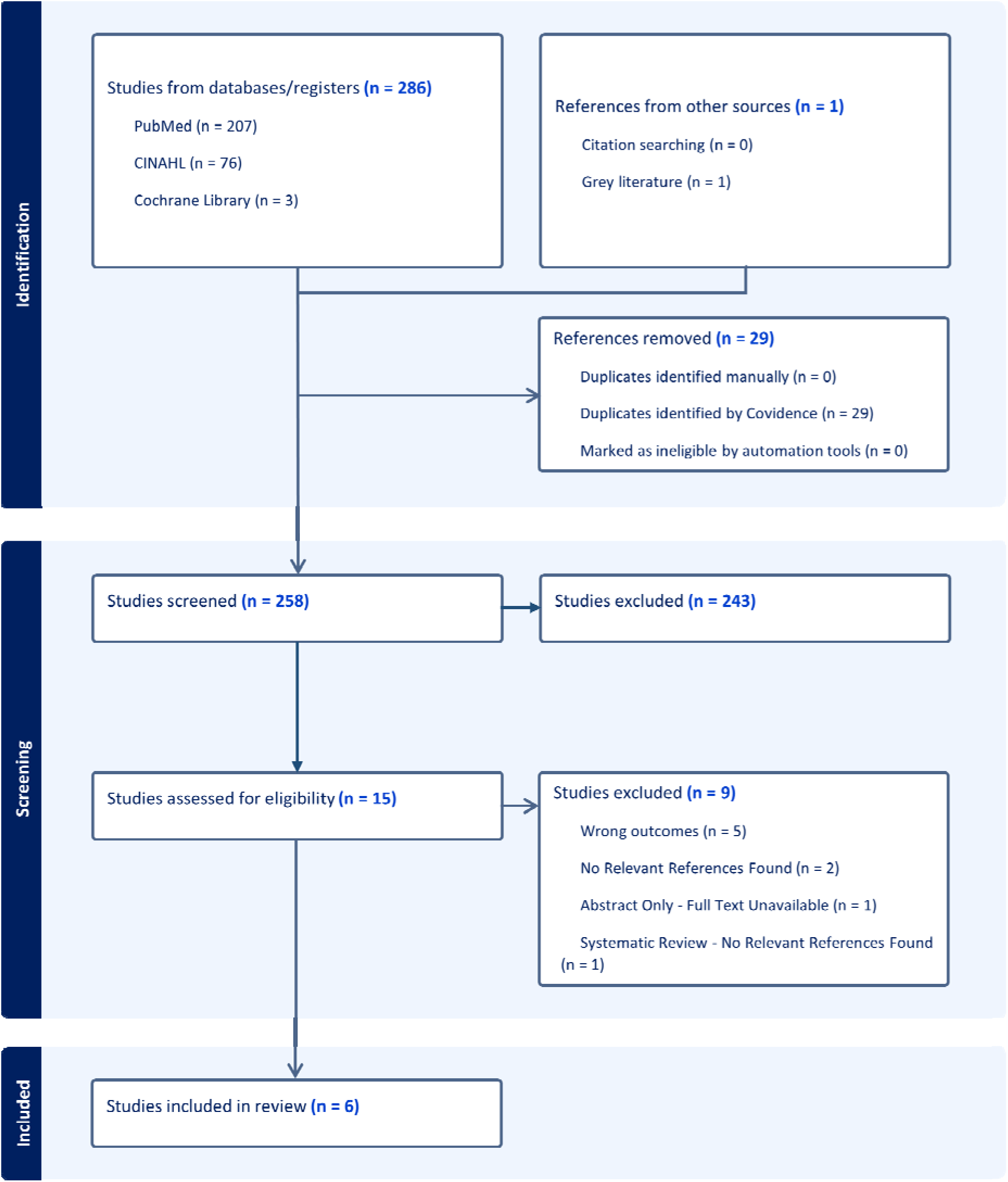
PRISMA Flow Diagram Showing the Process of Study Selection.

### Assessment of Data Quality

Critical appraisal or assessment of study quality was not carried out for reports included in this review. Thus, no study was excluded based on the quality of evidence if they met the review’s inclusion criteria. This was especially necessary since this is an area of public health in the UK where the current body of evidence is slowly or gradually gaining some research attention and where we expected that the breadth of the currently available evidence would be quite small.

### Data Extraction

A data extraction sheet was developed and pilot-tested on a sample of included studies. The final standardized data extraction sheet was designed to capture study characteristics, including Author(s), year of publication, study design, study setting, sample size, and participant demographics (age, ethnicity). The form also captured fibroid-related data such as prevalence, incidence, risk factors, symptoms, diagnostic methods, treatment approaches, outcomes, complications, quality of life, psychosocial impact, and healthcare utilisation. Key findings and conclusions related to fibroids in Caribbean and African women in the UK were also extracted. The extracted data was synthesised and presented narratively, accompanied by tables and figures. Results were organised as key themes identified during data extraction.

### Ethical Considerations

This rapid review did not involve collecting primary data from human participants, so ethical approval was not required.

## Results

A total of 287 citations were retrieved from academic (n=286) and grey literature (n=1) sources. No additional studies were retrieved following citation searching. Of the 287 citations retrieved, 29 were removed as duplicates. A further 243 citations were excluded from the remaining 258 for failing to meet the eligibility requirements for inclusion after title and abstract screening, leaving 15 articles for full text screening. Full text screening resulted in the exclusion of a further 9 citations, leaving six (6) articles at the end of the screening process for data extraction. The screening process is illustrated in the PRISMA flow diagram below (Figure 1).

### Study Characteristics

Studies were conducted between 2014 and 2020, primarily in the UK, with one joint study in the UK and the US. The studies employed various designs, including Case Reports, RCTs, and case-control, as well as either Retrospective or Prospective study designs. The sample size of participants, regardless of ethnicity, ranged from one in the case reports to 254 in the RCT. At least 275 African and/or Caribbean women participated across all studies, excluding the report by Edwards et al. (2019), in which no mention was made of the proportion of women of Black ethnicity among cases and controls. Excluding the case reports, there was no subgroup analysis of Black ethnicity; that is, no disaggregation of the data based on specific African or Caribbean ethnicities, such as “Black African,” “Black Caribbean,” “Mixed Black,” or “Mixed Caribbean,” was identified or reported (Table 1).

**Table 1.**
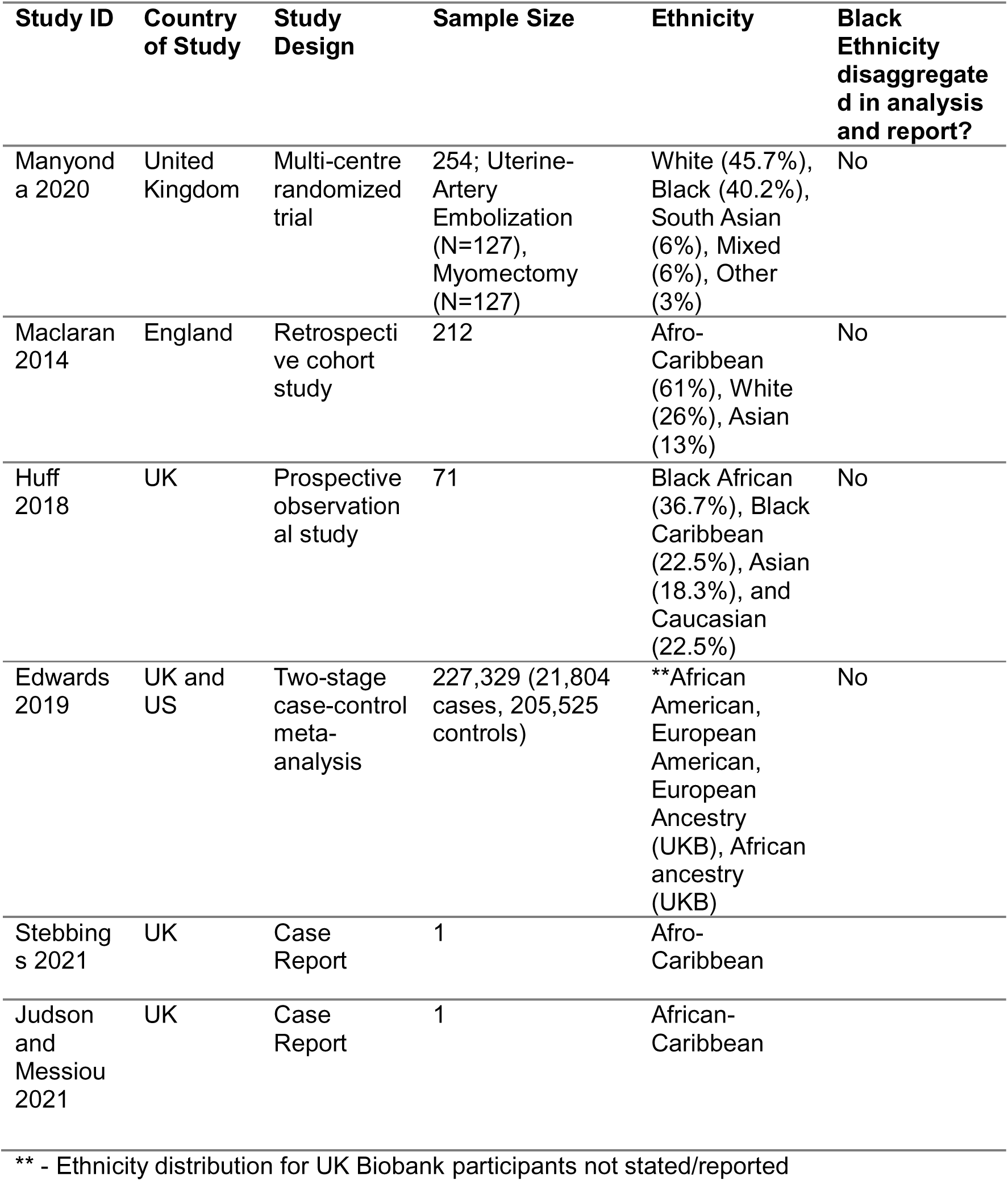
General Characteristics of Included Studies.

Edwards et al. (2019) reported the findings of a trans-ethnic genome-wide association study (GWAS) aimed at identifying genetic variants associated with the risk of uterine fibroids across European, African European, European American, and African American ancestry groups. The study utilized data from the electronic Medical Records and Genomics (eMERGE) network in the US and the UK Biobank. The analysis in this GWAS employed a two-stage approach: Stage I (discovery) included 9,446 cases and 67,048 controls, while Stage II (replication) comprised 12,358 cases and 138,477 controls, totalling 21,804 cases and 205,525 controls. Cases and controls were classified using non-cancer illness codes or ICD-10 codes for uterine leiomyomata. Approximately 14% of cases (1,325/9,446) and 3% of controls (1,927/67048) in Stage I were of African American or African ancestry from the UKB. The study identified 326 genome-wide significant variants across 11 loci, including three novel loci on chromosomes 1q24 (near *DNM3*), 16q12.1 (*HEATR3*), and 20q13.1 (*LOC105372640*), and confirmed eight previously reported loci. Notably, the *CDC42/WNT4* locus exhibited ancestry-specific effects, with opposite directions of association for African and European ancestry women; no significant associations were found at the locus for African American ancestry, compared to a significant increase in odds for the rs10917151 single nucleotide polymorphism (SNP) at the locus for European Africans. Genetically predicted gene expression analyses implicated genes such as *LUZP1, OBFC1, NUDT13,* and *HEATR3* in fibroid risk (Table 2).

**Table 2.**
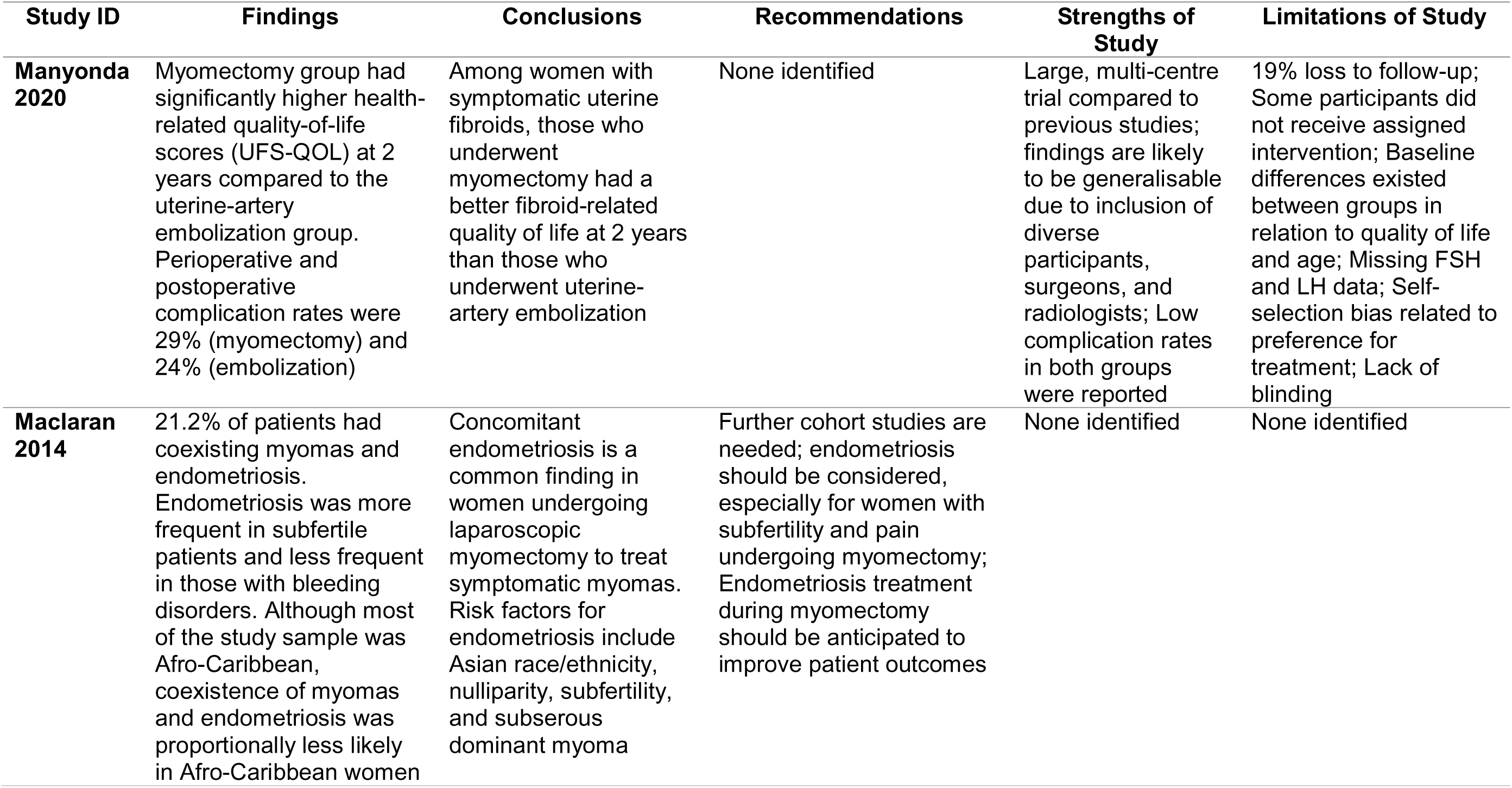
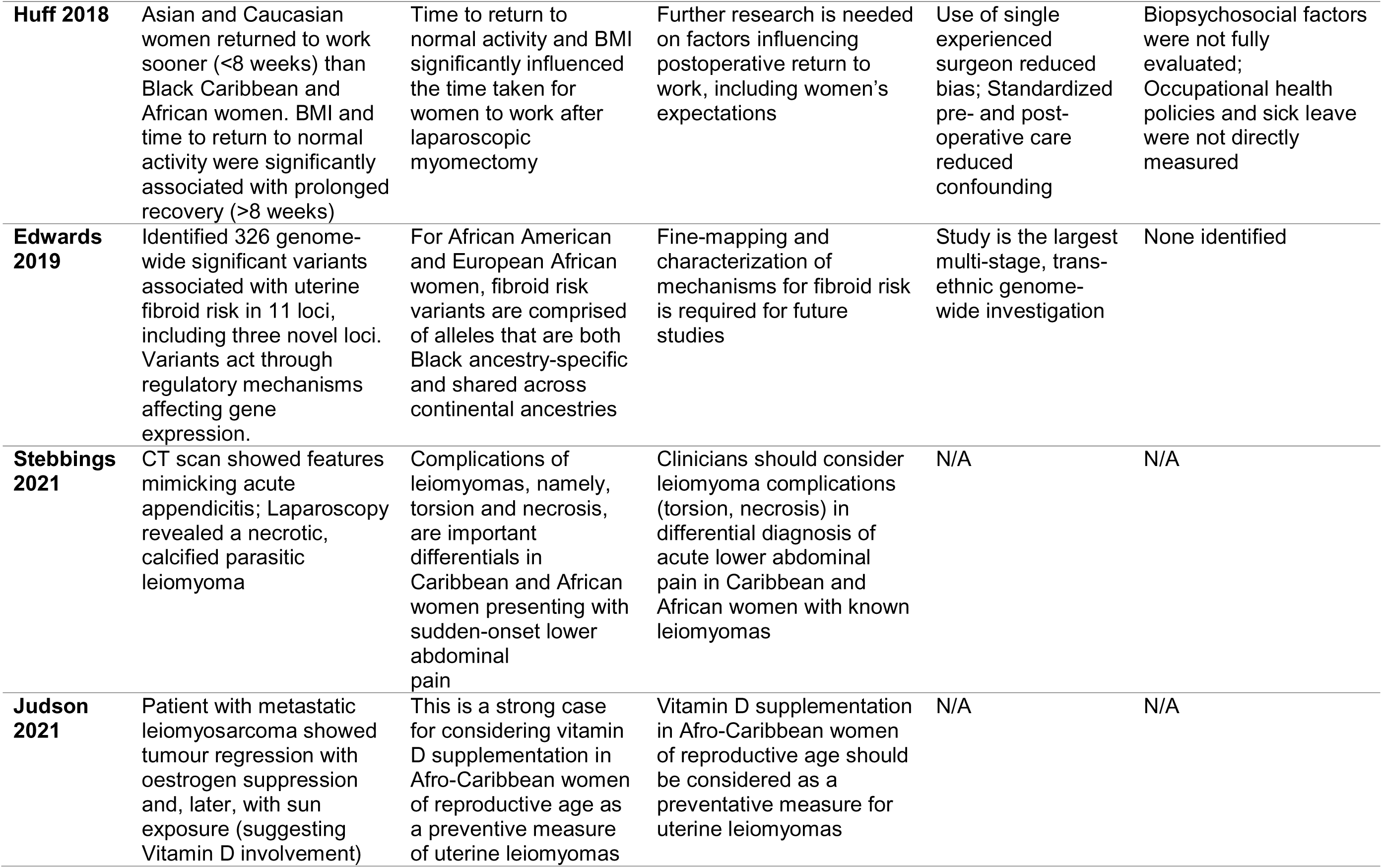
Summary of Findings of Included Studies.

| Study ID | Findings | Conclusions | Recommendations | Strengths of Study | Limitations of Study |
| --- | --- | --- | --- | --- | --- |
| **Manyonda 2020** | Myomectomy group had significantly higher health-related quality-of-life scores (UFS-QOL) at 2 years compared to the uterine-artery embolization group. Perioperative and postoperative complication rates were 29% (myomectomy) and 24% (embolization) | Among women with symptomatic uterine fibroids, those who underwent myomectomy had a better fibroid-related quality of life at 2 years than those who underwent uterine-artery embolization | None identified | Large, multi-centre trial compared to previous studies; findings are likely to be generalisable due to inclusion of diverse participants, surgeons, and radiologists; Low complication rates in both groups were reported | 19% loss to follow-up; Some participants did not receive assigned intervention; Baseline differences existed between groups in relation to quality of life and age; Missing FSH and LH data; Self-selection bias related to preference for treatment; Lack of blinding |
| **Maclaran 2014** | 21.2% of patients had coexisting myomas and endometriosis. Endometriosis was more frequent in subfertile patients and less frequent in those with bleeding disorders. Although most of the study sample was Afro-Caribbean, coexistence of myomas and endometriosis was proportionally less likely in Afro-Caribbean women | Concomitant endometriosis is a common finding in women undergoing laparoscopic myomectomy to treat symptomatic myomas. Risk factors for endometriosis include Asian race/ethnicity, nulliparity, subfertility, and subserous dominant myoma | Further cohort studies are needed; endometriosis should be considered, especially for women with subfertility and pain undergoing myomectomy; Endometriosis treatment during myomectomy should be anticipated to improve patient outcomes | None identified | None identified |
| **Huff 2018** | Asian and Caucasian women returned to work sooner (<8 weeks) than Black Caribbean and African women. BMI and time to return to normal activity were significantly associated with prolonged recovery (>8 weeks) | Time to return to normal activity and BMI significantly influenced the time taken for women to work after laparoscopic myomectomy | Further research is needed on factors influencing postoperative return to work, including women's expectations | Use of single experienced surgeon reduced bias; Standardized pre- and post-operative care reduced confounding | Biopsychosocial factors were not fully evaluated; Occupational health policies and sick leave were not directly measured |
| **Edwards 2019** | Identified 326 genome-wide significant variants associated with uterine fibroid risk in 11 loci, including three novel loci. Variants act through regulatory mechanisms affecting gene expression. | For African American and European African women, fibroid risk variants are comprised of alleles that are both Black ancestry-specific and shared across continental ancestries | Fine-mapping and characterization of mechanisms for fibroid risk is required for future studies | Study is the largest multi-stage, trans-ethnic genome-wide investigation | None identified |
| **Stebbings 2021** | CT scan showed features mimicking acute appendicitis; Laparoscopy revealed a necrotic, calcified parasitic leiomyoma | Complications of leiomyomas, namely, torsion and necrosis, are important differentials in Caribbean and African women presenting with sudden-onset lower abdominal pain | Clinicians should consider leiomyoma complications (torsion, necrosis) in differential diagnosis of acute lower abdominal pain in Caribbean and African women with known leiomyomas | N/A | N/A |
| **Judson 2021** | Patient with metastatic leiomyosarcoma showed tumour regression with oestrogen suppression and, later, with sun exposure (suggesting Vitamin D involvement) | This is a strong case for considering vitamin D supplementation in Afro-Caribbean women of reproductive age as a preventative measure of uterine leiomyomas | Vitamin D supplementation in Afro-Caribbean women of reproductive age should be considered as a preventative measure for uterine leiomyomas | N/A | N/A |

The study by Manyonda et al. (2020) reported on the findings of the study “Treating Fibroids with Either Embolisation or Myomectomy to Measure the Effect on Quality of Life Among Women Wishing to Avoid Hysterectomy (FEMME) trial, a multi-centre, randomised, open-label study conducted at 29 hospitals in the United Kingdom”. Their study aimed to separately compare the effectiveness of myomectomy and uterine-artery embolisation (UAE) in women with symptomatic uterine fibroids who wished to preserve their uterus. The study utilised a 1:1 randomisation of 254 premenopausal women older than 18 years of age. Eligibility for this study included not being pregnant and being symptomatic for uterine fibroids. Myomectomy procedures included open abdominal, laparoscopic, or hysteroscopic approaches. The primary outcome was fibroid-related quality of life at 2 years, assessed using the Uterine Fibroid Symptom and Quality of Life (UFS-QOL) questionnaire. Of the participants, 48 (38%) in the UAE group and 54 (43%) in the myomectomy group identified as Black. The intention-to-treat analysis of 206 women (81%) revealed that myomectomy resulted in a significantly better quality of life score (84.6 ± 21.5) compared to UAE (80.0 ± 22.0), with a mean adjusted difference of 8.0 points (95% CI (1.8 to 14.1), P=0.01). Perioperative and postoperative complications were similar between groups (29% myomectomy vs. 24% UAE) (Table 2).

The study by Maclaran et al., (2014) retrospectively investigated the prevalence of coexisting uterine myomas and endometriosis in women undergoing laparoscopic myomectomy. It explored associated risk factors and surgical implications among a cohort of women at a tertiary referral centre in London, England. Data were collected from 212 women who underwent laparoscopic myomectomy for symptomatic uterine myomas between 2005 and 2013. The presence of endometriosis was confirmed either visually during laparoscopy or by histopathology. The study population’s mean age was 38.1 years, with a mean BMI of 26.7. The cohort was predominantly Afro-Caribbean (61%), with 26% white and 13% Asian participants. Analysis revealed that 21.2% (45 women) had coexisting endometriosis. This coexistence was significantly more common in women with subfertility (44% vs. 25.7%) and less common in those with bleeding disorders (20% vs. 45%). While parity, location of myomas and race were related to an increased risk of the coexistence of both conditions, the size or number of myomas was not. Afro-Caribbean women were proportionally less likely to have coexisting myomas and endometriosis (OR=0.41, 95% CI (0.21 to 0.81), p=0.01). Of those with endometriosis, 42% underwent surgical treatment for it during myomectomy. Patients with endometriosis also had significantly more ovarian cystectomies. However, operative time and blood loss were similar between groups with and without concurrent endometriosis (Table 2).

Huff et al. (2018) reported on a prospective observational study conducted at Whipps Cross University Hospital in London, aimed at evaluating the time taken for women to return to work after laparoscopic myomectomy and identifying factors that prolong recovery beyond 8 weeks. The study involved 94 women undergoing the procedure performed by a single surgeon between January 2012 and March 2015. Data were collected through a validated return-to-work questionnaire completed postoperatively at 3 months. Of the 71 women who completed the questionnaire, 42 (59.1%) identified as Black African or Black Caribbean ethnicity (26 (36.7%) being Black African and 16(22.5%) Black Caribbean). The findings revealed that a significant proportion of women (39.4%) took longer than 8 weeks to return to work. Body Mass Index (BMI) and time to return to normal activity were the only factors significantly associated with a prolonged return to work. Notably, a significantly lower proportion of Black Caribbean and African women (19 out of 42) returned to work within 8 weeks compared to other ethnicities in the study (Asian and Caucasian women), with a significantly higher mean number of fibroids removed from Black Caribbean and African women potentially contributing to this difference (Table 2).

The case report by Stebbings et al. in 2021 aimed to highlight a rare but significant differential diagnosis of a torsed parasitic leiomyoma (PL) in an Afro-Caribbean woman presenting with lower abdominal pain, initially thought to be related to acute appendicitis. The study was conducted at Lewisham and Greenwich NHS Trust and Barts Health NHS Trust in London. It involved a 39-year-old multiparous Afro-Caribbean woman who presented to the emergency department with sudden-onset, severe right iliac fossa pain. The methods used included physical examination, blood tests (which revealed mildly elevated inflammatory markers), urine tests, and an emergency out-of-hours CT scan of the abdomen and pelvis. The scan, however, showed significantly calcified uterine leiomyomas in addition to suspected appendicitis. Due to the mixed presentation, the patient underwent an emergency diagnostic laparoscopy, which found her appendix to be macroscopically normal. A necrotic, heavily calcified PL was discovered detached from the uterus. The PL was successfully removed laparoscopically, and histological examination of the excised tissue confirmed calcification consistent with an infarcted/necrotic leiomyoma. The study posited that while a torsed PL is rare, its symptoms can mimic acute appendicitis and should be considered in the differential diagnosis of women, particularly those of Black ethnicity presenting with sudden-onset severe lower abdominal pain, especially those with a known history of uterine leiomyomas (Table 2).

The case report by Judson and Messiou in 2021 aimed to highlight the potential link between vitamin D deficiency and the pathogenesis of leiomyoma and intravascular leiomyomatosis among women of Afro-Caribbean ancestry. The patient, a 36-year-old woman, presented in 2006 at The Royal Marsden NHS Foundation Trust in London with multiple uterine leiomyomata, a tumour invading the inferior vena cava (IVC), and pulmonary metastases. The methods involved surgical removal of the intravascular tumour, and following recurrence, imaging techniques such as CT and MRI scans were employed to monitor disease progression. Initially, the patient was treated with oestrogen suppression therapy (goserelin and letrozole), but upon follow-up after extended exposure to sunlight in a tropical environment, she reported an improvement in her symptoms. The authors suggest that their findings indicate a potential benefit of vitamin D, through sunlight exposure and supplementation, in stabilising and reducing the size of uterine leiomyomas and intravascular leiomyomatosis among women of Caribbean and African ancestry. They also propose that this finding suggests a role for low levels of vitamin D in the pathogenesis of uterine leiomyomas in women of Caribbean and African ethnicity (Table 2).

## Discussion

The purpose of this review was to scope and examine the evidence in the UK regarding uterine fibroids among women of African and/or Caribbean ancestry. By utilizing a systematic approach to examine both peer-reviewed and grey literature, this review identified a limited number of studies (six studies) documenting fibroids among Black women in the UK. The review also found evidence related to the genetics of fibroids, treatment options, and their connections to quality of life, recovery time, complications, and severe presentations of the condition, as well as the potential of vitamin D supplementation as a protective factor. The following sections discuss these findings in more detail.

### 1. The Genetics of Fibroids among Women of African/Caribbean Ethnicity in the UK

Generally, experts agree that there is significant evidence pointing to a heritable component in fibroid development. Twin studies have estimated that the heritability of fibroid genes may be as high as 70%, suggesting that genetic factors could play a major role (Baird et al., 2002). Furthermore, familial aggregation studies consistently show a higher risk of fibroids among women who have immediate family members diagnosed with the condition (Ligon and Morton, 2000).

Besides the evidence supporting the heritability of fibroids, there is a consensus that multiple genes may contribute to the development of this condition. The development of fibroids is likely polygenic, involving several susceptibility gene loci with varying effect sizes. This is exemplified by the absence of evidence for traditional Mendelian inheritance patterns and the identification of numerous associated genetic variants through genome-wide association studies (GWAS) (Wise et al. 2015).

Another argument used to support the genetic basis of fibroids is the observation of the development of the condition, particularly among women of specific ethnicities. African ancestry is a significant risk factor for fibroids, with a higher prevalence, earlier age of onset, and more severe disease found in this demographic compared to other racial groups. Although socioeconomic and environmental factors likely contribute, genetic differences have also been implicated. For example, Stewart et al. (2016) demonstrated that African American women were two to three times more likely to be diagnosed with fibroids than Caucasian women. This disparity suggests potential ancestry-specific risk alleles.

In this review, we have provided evidence in support of the genetic basis of fibroid development and diagnosis, particularly among women of Black ethnic origins or ethnicity. We have also shown that multiple genes could be involved among these women, with the large GWAS study by Edwards et al. in 2019 showing concordance between the US and UK samples in the loci of interest (11 loci found in all; 8 shared and 3 new/additional loci).

We did not find published literature on the familial inheritance of fibroids among Caribbean and African women living in the UK, suggesting that more evidence is needed to understand how familial gene inheritance may increase or transfer fibroid risk/ susceptibility.

When considering the evidence for the genetic basis of fibroids, there are areas of considerable debate that should be addressed in any future studies on fibroids among Black women in the UK. For example, while GWAS has identified numerous single nucleotide polymorphisms (SNPs) linked to the development of the disease, pinpointing the specific causal pathways and their functional roles in fibroid pathogenesis remains a challenge (Cha et al. 2018). Additionally, there is a potential interaction between genes and the environment, where many confounders may exist (e.g., diet and exposure to endocrine-disrupting chemicals). This further illustrates how poorly understood fibroid progression is, especially for Black women living in the UK. Emerging evidence also suggests that epigenetic modifications, including DNA methylation and histone modifications, may play a role in fibroid development and contribute to racial disparities (Asad et al. 2012). However, the extent and specific mechanisms of epigenetic involvement remain unclear.

In summary, although a strong genetic contribution to uterine fibroid development is well-established, the intricate details of the underlying genetic architecture remain to be fully elucidated, particularly among Black women in the UK. Further research, including fine-mapping of GWAS loci, functional studies of candidate genes, and investigations into gene-environment and epigenetic interactions, is crucial for achieving a more comprehensive understanding of the complex genetic basis of this disease and for developing more targeted prevention and treatment strategies.

### 2. Treatment Options and their Relationships with the Quality of Life and Recovery Time

There are various treatment options for uterine fibroids currently available, ranging from expectant management to pharmacological/medical and surgical interventions. Treatment decisions are typically individualised and based on factors such as symptom severity, patient preference, the desire for future pregnancies, and the size, number, and location of fibroid tumours.

For asymptomatic fibroids, expectant management (observation with regular monitoring) is the preferred approach (ACOG, 2018). However, pharmacologic and other medical therapies, including gonadotropin-releasing hormone (GnRH) agonists, selective progesterone receptor modulators (SPRMs), and tranexamic acid, are also used to effectively reduce fibroid size and alleviate symptoms such as heavy menstrual bleeding (Donnez et al. 2002). These therapies, however, often provide temporary relief and can be associated with side effects that limit their long-term use.

Definitive treatment is typically achieved through the surgical removal of the tumour. Currently, surgical options include myomectomy (which involves the removal of fibroids while preserving the uterus) and hysterectomy (which entails the removal of the uterus) (ACOG, 2018). However, to reduce or eliminate the possibility of recurrence, hysterectomy is generally considered the most definitive treatment option. For Caribbean and African women in the UK, evidence from this review, such as Huff et al. 2018, indicates a notable preference for preserving the uterus, particularly among those women interested in future pregnancies.

Despite the promise of treatment, there are still areas where consensus is needed, and it is crucial to consider these aspects as they are relevant to any discussion about the treatment of fibroids among women of African or Caribbean descent living in the UK. These areas include the optimal approach to myomectomy, the long-term effects of Uterine Artery Embolisation (UAE), quality of life outcomes after surgery, and the recovery time for patients post-surgery.

Currently, there are three surgical approaches to myomectomy: laparoscopy, hysteroscopy, and abdominal myomectomy. However, the choice between laparoscopic, hysteroscopic, and abdominal myomectomy depends on the size, number, and location of the fibroid. Each approach has its own advantages and disadvantages concerning recovery time, complications, and long-term outcomes.

For Black women in the UK this is critical, as post-surgery outcomes or complications for invasive approaches have been reported to be severe (Majak et al., 2012). However, minimally invasive approaches, such as laparoscopy and hysteroscopy-which may be the more preferred option for Black women- are worth exploring. These methods are linked to shorter hospital stays, less postoperative pain, and a faster return to normal activities compared to the more invasive abdominal myomectomy (Laughlin-Tommaso et al., 2013). The feasibility and effectiveness of minimally invasive techniques could, however, be limited by fibroid characteristics, which, as previously mentioned, are more severe among Caribbean and African women.

Minimally invasive approaches, such as UAE, may effectively reduce fibroid size and symptoms among Black women in the UK; however, concerns persist about its overall impact on future fertility and potential long-term complications (Mara et al. 2008).

Although many studies report improvements in quality of life following fibroid treatment, few have made direct comparisons of different treatment modalities using standardized quality-of-life measures. For example, in this review, we found only one study, specifically Manyonda et al. 2020, where a standard quality-of-life questionnaire was administered to patients who preferred less invasive treatment options, such as UAE or Myomectomy, outside of hysterectomy. More research is needed to directly compare the long-term quality-of-life outcomes associated with various fibroid treatment options (Flyckt et al. 2017). Additionally, factors beyond symptom relief, including sexual function, body image, and psychological well-being, must be considered, especially among Caribbean and African women in the UK.

Recovery time after fibroid treatment varies based on the chosen procedure, individual patient characteristics such as the number and size of fibroids, and the definition of recovery. Invasive procedures generally lead to longer recovery times (reference). This review provides evidence from Huff et al.’s study in 2018, which suggests that Caribbean and African women who present with significantly more fibroids or more aggressive forms of the tumour (compared to women of other ethnicities) are likely to spend longer in recovery and take more time to return to work. Among Caribbean and African women in the UK, further research examining the standardization of reporting recovery times and a comprehensive assessment of factors influencing recovery is needed to better evaluate the most effective treatment modalities.

### 3. Complications and Extreme Presentations of Symptoms

There is a consensus that the common complications associated with uterine fibroids include heavy blood loss (menorrhagia), pelvic pain, and reproductive issues. Menorrhagia often leads to anaemia and a reduced quality of life (Stewart, 2001); pelvic pain is commonly associated with large tumours due to their mass effect on surrounding organs (Wallach et al. 1998); and typical reproductive issues include infertility and miscarriage (Pritts et al. 2009). In terms of pregnancy outcomes,

These complications are notably frequent, particularly among Caribbean and African women who report larger tumour sizes and a comparatively higher number of fibroids. In this review, we have identified evidence that confirms or highlights some of these complications, such as pelvic pain (which was initially mistaken for appendicitis), in the case report by Stebbings et al. (2021).

Depending on the location-submucosal, intramural, or subserosal fibroids may also be linked to pregnancy complications such as infertility. While the association between submucosal fibroids and infertility is well-established, the impact of other types (intramural and subserosal) on fertility remains less clear (Saridogan et al. 2016). Some studies even suggest that these types may negatively affect conception rates, but the evidence is still inconclusive (Saridogan et al. 2016). Further research is needed to understand how the location and severity of fibroids could impact infertility or miscarriage, particularly among Africans and Caribbeans in the UK. Other complications that may warrant further investigation include birth delivery by caesarean section and urinary incontinence, where the evidence in the broader literature-although not specific to Black ethnicity in the UK- also remains unclear.

Rarely, fibroid complications can be severe (Parker, 2007). In this review, we report evidence supporting this finding, such as in the study by Stebbings et al. (2021), where the fibroid tumour was found to be parasitic and calcified, leading to lower abdominal pain. We also present evidence of cases where benign fibroid tumour cells could potentially become malignant and spread intravascularly to other areas of the body (Judson and Messiou, 2021). It remains unclear exactly how these changes could occur, and more research is necessary to understand the molecular and physiological basis for these changes to improve care for women of Caribbean and African ethnicity living in the UK.

For Black women in the UK, the complications of fibroids could also potentially affect birth outcomes. Black women with fibroids have demonstrated a higher rate of preterm birth compared to those without fibroids, and this risk is particularly higher for earlier gestational ages (Landman et al., 2022). This is particularly relevant for Black women, who are also at a higher risk for fibroids and may face compounded risks during pregnancy (Landman et al., 2022).

### 4. Hypovitaminosis D as a Risk Factor / Vitamin D Supplementation as a potential Protective Factor

The relationship between vitamin D status and the risk and development of uterine fibroids is an area of ongoing research. While no definitive consensus exists, multiple observational studies have reported an inverse association between vitamin D levels and the risk of uterine fibroids (Vitale et al. 2021). Women with lower serum 25-hydroxyvitamin D [25(OH)D] levels, indicating vitamin D deficiency or insufficiency (hypovitaminosis D), have a higher risk of developing fibroids (Bae et al. 2018). Although observational studies suggest an association, they do not establish causality. It remains unclear whether vitamin D deficiency directly contributes to fibroid development or if the observed association is due to confounding factors or reverse causality (Vitale et al. 2021).

The potential biological mechanisms linking vitamin D to fibroid pathogenesis are still under investigation. However, the proposed mechanisms include regulating cell proliferation and apoptosis, modulating inflammation and the extracellular matrix, and regulating oestrogen signalling.

Vitamin D has anti-proliferative and pro-apoptotic effects in various cell types, including smooth muscle cells. The disruption of these pathways due to vitamin D deficiency may contribute to fibroid growth (Halder et al. 2011). Additionally, vitamin D possesses anti-inflammatory properties and may influence the production and remodelling of the extracellular matrix. Furthermore, vitamin D may interact with oestrogen signalling pathways, which play a crucial role in fibroid growth. However, the specifics of this interaction remain unclear.

Due to the lack of conclusive evidence for a causal relationship, there are currently no clinical recommendations for vitamin D supplementation as a preventive or treatment strategy for uterine fibroids (Halder et al. 2018), both generally and specifically for Caribbean and African women in the UK. Well-designed randomised controlled trials are necessary to determine whether vitamin D supplementation can effectively reduce the risk or growth of fibroids among women of Black ethnic origin (especially those in the UK). Research should also identify the optimal dosage, timing of community entry, and the critical age for intervention or supplementation. Given that the currently available evidence suggesting that vitamin D might serve as an environmental protective factor against the development of fibroids among Caribbean and African women in the UK, the relationship between vitamin D status and genetic predisposition needs further investigation.

Available evidence also suggests that a high prevalence of vitamin D deficiency (independent of the presence of fibroids) also contributes to adverse pregnancy outcomes among women of African ethnic origin/descent (Ayamah et al. 2025). This potentially presents a double burden of deficiency in which the pathway linking vitamin D deficiency to fibroid development and subsequent adverse pregnancy outcomes may be amplified in this population. At the population level, this could contribute to persistent racial disparities in maternal and neonatal health, including higher rates of preterm birth (Landman et al. 2022).

### Strengths of Study

The primary strength of this review lies in its systematic approach to identifying studies, extracting data, and analysing/synthesizing findings. Furthermore, while the included studies are limited in number, they provide diverse perspectives through a range of study designs. This diversity allows for a multifaceted understanding of the available literature, encompassing genetic factors, treatment effectiveness, work and quality of life, as well as clinical presentations.

### Limitations of Study

Despite its strengths, this review has limitations related to its methodology and evidence base. As a rapid review, the literature search has limitations compared to a systematic review. Specifically, this limitation pertains to the number of relevant academic databases searched. Although additional grey literature searches were conducted, it is possible that some pertinent studies, particularly those not indexed in major databases or easily accessible online, may have been overlooked.

Critical appraisal or quality assessment of included studies was not conducted for this study. Although this allowed for the inclusion of the small number of studies found, the methodological rigor and potential biases within the primary studies were not independently assessed. Consequently, any conclusions drawn from this review should be interpreted with caution.

Furthermore, the limited number of studies included (six) underscores the scarcity of research on uterine fibroids among Caribbean and African women in the UK. This scarcity also restricts the generalizability of the review’s findings. The included studies also reported small sample sizes, potential selection bias, and an absence of disaggregated data for specific Caribbean and African sub-groups.

Critically, this review did not identify any qualitative studies that focused on the rich lived experiences of Caribbean and African women in the UK who had either been diagnosed with or were living with fibroids. This lack of qualitative studies imposes additional limitations into gaining further insight and philosophical depth for some of the quantitative findings in this review.

## Recommendations for Policy, Practice, and Research

Based on the findings of this rapid review, we have proposed several key recommendations for policy, practice, and future research.

In terms of policy, there is a clear need to acknowledge and address the disproportionate burden of uterine fibroids faced by this population within national health strategies. Policy initiatives should prioritise increasing awareness of uterine fibroids among Caribbean and African women, ensuring culturally sensitive and accessible information is available regarding risk factors, symptoms, and treatment options. This could involve targeted public health campaigns in community settings, collaboration with community organisations and leaders, and the development of culturally tailored educational materials in various formats and languages. Furthermore, policies should support the development of healthcare services that are responsive to the specific needs of Caribbean and African women with fibroids. This includes ensuring equitable access to diagnostic services, a range of treatment options, including minimally invasive approaches, and specialist care when needed. Policymakers should also consider initiatives to address wider social determinants of health that may contribute to fibroid risk and outcomes in this population, such as socioeconomic factors, dietary patterns, and vitamin D deficiency.

In terms of clinical practice, the review highlights the importance of culturally competent and patient-centred care for Caribbean and African women with uterine fibroids. Healthcare professionals should be aware of the higher prevalence, earlier onset, and potentially more severe presentations of fibroids in this population. Clinical guidelines and protocols need to be adapted to address these disparities and ensure that Caribbean and African women receive timely and appropriate diagnosis and treatment. Clinicians should engage in open and respectful communication with patients, exploring their preferences, concerns, and cultural beliefs regarding treatment options, particularly those related to uterine preservation and fertility. Given the evidence suggesting a preference for uterine preservation in this population, clinicians should ensure that minimally invasive options like myomectomy and UAE are readily discussed and offered when clinically appropriate. Furthermore, healthcare practitioners should be vigilant in considering differential diagnoses, as highlighted by the case report of a torsed parasitic leiomyoma, especially in Caribbean and African women presenting with atypical abdominal pain. Routine assessment of vitamin D status and consideration of supplementation, particularly in women at risk for deficiency, could also be integrated into clinical practice. However, further research is needed to confirm its preventive and therapeutic role.

In terms of future research, this review underscores the urgent need for more robust studies on uterine fibroids among Caribbean and African women in the UK. There is a critical need for large-scale epidemiological studies to accurately determine the prevalence, incidence, and risk factors for fibroids in this population, with a detailed disaggregation of ethnicity data to understand variations within Caribbean and African sub-groups. Longitudinal studies are essential to investigate the natural history of fibroids in this population, including disease progression, treatment outcomes, and long-term quality of life impacts. Comparative effectiveness studies are required to evaluate the optimal treatment approaches for Caribbean and African women with fibroids, considering both clinical outcomes and patient-reported outcomes (quality of life, recovery time, and satisfaction). Further research is also necessary to explore the genetic and epigenetic mechanisms and interactions underpinning fibroid disparities. This could include fine-mapping of GWAS loci and investigations into gene-environment interactions in this population. Well-designed randomized controlled trials are crucial to investigate the potential role of vitamin D supplementation in preventing or treating uterine fibroids in Caribbean and African women in the UK, determining optimal dosage, timing, and community entry points for intervention. These trials may also be extended to evaluate the benefits of vitamin D supplementation on improving birth outcomes, including the high prevalence of pre-term births and miscarriages among Black women in the UK. Finally, we recommend that studies capturing the lived experiences of Black women in the UK who have been diagnosed or living with fibroids should be conducted in order to gain further depth and insights on any quantitative findings.

## Conclusion

This rapid review, although constrained by the limited availability of existing evidence, offers a valuable overview of the current state of knowledge about uterine fibroids among Caribbean and African women in the UK. The review confirms the disproportionate burden of fibroids in this population, emphasizing the high prevalence, early onset, and severity of symptoms. It also highlights the significance of genetic predisposition and environmental factors in contributing to fibroid development within this group. While treatment options exist, the review indicates areas where consensus remains necessary, particularly concerning optimal surgical approaches, long-term effects of UAE, and quality-of-life outcomes. Furthermore, the review points to the potential of vitamin D as a protective factor.

## Data Availability

All data produced in the present study are available upon reasonable request to the authors.

## Notes

### Competing Interest Statement

The authors have declared no competing interest.

### Funding Statement

This study did not receive any funding.

