## Supplementary Material 1 for "Uterine Fibroids Among Caribbean and African Women in the UK: A Rapid Scoping Review"

**Supplementary Material 1: Literature Search Strategy and Records Retrieved**

**PubMed (n=207)**

| Search number | Query | Results |
| --- | --- | --- |
| 16 | ((((((((((((((("Aberdeen"[Affiliation] OR "Armagh"[Affiliation] OR "Bangor"[Affiliation] OR "Bangor"[Affiliation] OR "Bath"[Affiliation] OR "Belfast"[Affiliation] OR "Birmingham"[Affiliation] OR "Bradford"[Affiliation] OR "Brighton & Hove"[Affiliation] OR "Bristol"[Affiliation] OR "Cambridge"[Affiliation] OR "Canterbury"[Affiliation] OR "Cardiff"[Affiliation] OR "Carlisle"[Affiliation] OR "Chelmsford"[Affiliation] OR "Chester"[Affiliation] OR "Chichester"[Affiliation] OR "Colchester"[Affiliation] OR "Coventry"[Affiliation] OR "Derby"[Affiliation] OR "Doncaster"[Affiliation] OR "Dundee"[Affiliation] OR "Dunfermline"[Affiliation] OR "Durham"[Affiliation] OR "Edinburgh"[Affiliation] OR "Ely"[Affiliation] OR "England"[Affiliation] OR "Exeter"[Affiliation] OR "Glasgow"[Affiliation] OR "Gloucester"[Affiliation] OR "Hereford"[Affiliation] OR "Inverness"[Affiliation] OR "Kingston-Upon-Hull"[Affiliation] OR "Lancaster"[Affiliation] OR "Leeds"[Affiliation] OR "Leicester"[Affiliation] OR "Lichfield"[Affiliation] OR "Lincoln"[Affiliation] OR "Lisburn"[Affiliation] OR "Liverpool"[Affiliation] OR "London"[Affiliation] OR "Londonderry"[Affiliation] OR "Manchester"[Affiliation] OR "Milton Keynes"[Affiliation] OR "Newcastle-Upon-Tyne"[Affiliation] OR "Newport"[Affiliation] OR "Newry"[Affiliation] OR "Northern Ireland"[Affiliation] OR "Norwich"[Affiliation] OR "Nottingham"[Affiliation] OR "Oxford"[Affiliation] OR "Perth"[Affiliation] OR "Peterborough"[Affiliation] OR "Plymouth"[Affiliation] OR "Portsmouth"[Affiliation] OR "Preston"[Affiliation] OR "Ripon"[Affiliation] OR "Salford"[Affiliation] OR "Salisbury"[Affiliation] OR "Scotland"[Affiliation] OR "Sheffield"[Affiliation] OR "Southampton"[Affiliation] OR "Southend-on-Sea"[Affiliation] OR "St Albans"[Affiliation] OR "St Asaph"[Affiliation] OR "St Davids"[Affiliation] OR "Stirling"[Affiliation] OR "Stoke on Trent"[Affiliation] OR "Sunderland"[Affiliation] OR "Swansea"[Affiliation] OR "Truro"[Affiliation] OR "United Kingdom"[Affiliation] OR "Wakefield"[Affiliation] OR "Wales"[Affiliation] OR "Wells"[Affiliation] OR "Westminster"[Affiliation] OR "Winchester"[Affiliation] OR "Wolverhampton"[Affiliation] OR "Worcester"[Affiliation] OR "Wrexham"[Affiliation] OR "York"[Affiliation]) OR ("Aberdeen"[Title/Abstract] OR "Armagh"[Title/Abstract] OR "Bangor"[Title/Abstract] OR "Bath"[Title/Abstract] OR "Belfast"[Title/Abstract] OR "Birmingham"[Title/Abstract] OR "Bradford"[Title/Abstract] OR "Brighton hove"[Title/Abstract] OR "Bristol"[Title/Abstract] OR "Cambridge"[Title/Abstract] OR "Canterbury"[Title/Abstract] OR "Cardiff"[Title/Abstract] OR "Carlisle"[Title/Abstract] OR "Chelmsford"[Title/Abstract] OR "Chester"[Title/Abstract] OR "Chichester"[Title/Abstract] OR "Colchester"[Title/Abstract] OR "Coventry"[Title/Abstract] OR "Derby"[Title/Abstract] OR "Doncaster"[Title/Abstract] OR "Dundee"[Title/Abstract] OR "Dunfermline"[Title/Abstract] OR "Durham"[Title/Abstract] OR "Edinburgh"[Title/Abstract] OR "Ely"[Title/Abstract] OR "England"[Title/Abstract] OR "Exeter"[Title/Abstract] OR "Glasgow"[Title/Abstract] OR "Gloucester"[Title/Abstract] OR "Hereford"[Title/Abstract] OR "Inverness"[Title/Abstract] OR "Kingston-Upon-Hull"[Title/Abstract] OR "Lancaster"[Title/Abstract] OR "Leeds"[Title/Abstract] OR "Leicester"[Title/Abstract] OR "Lichfield"[Title/Abstract] OR "Lincoln"[Title/Abstract] OR "Lisburn"[Title/Abstract] OR "Liverpool"[Title/Abstract] OR "London"[Title/Abstract] OR "Londonderry"[Title/Abstract] OR "Manchester"[Title/Abstract] OR "Milton Keynes"[Title/Abstract] OR "Newcastle-Upon-Tyne"[Title/Abstract] OR "Newport"[Title/Abstract] OR "Newry"[Title/Abstract] OR "Northern Ireland"[Title/Abstract] OR "Norwich"[Title/Abstract] OR "Nottingham"[Title/Abstract] OR "Oxford"[Title/Abstract] OR "Perth"[Title/Abstract] OR "Peterborough"[Title/Abstract] OR "Plymouth"[Title/Abstract] OR "Portsmouth"[Title/Abstract] OR "Preston"[Title/Abstract] OR "Ripon"[Title/Abstract] OR "Salford"[Title/Abstract] OR "Salisbury"[Title/Abstract] OR "Scotland"[Title/Abstract] OR "Sheffield"[Title/Abstract] OR "Southampton"[Title/Abstract] OR "Southend-on-Sea"[Title/Abstract] OR "St Albans"[Title/Abstract] OR "St Davids"[Title/Abstract] OR "Stirling"[Title/Abstract] OR "Stoke on Trent"[Title/Abstract] OR "Sunderland"[Title/Abstract] OR "Swansea"[Title/Abstract] OR "Truro"[Title/Abstract] OR "United Kingdom"[Title/Abstract] OR "Wakefield"[Title/Abstract] OR "Wales"[Title/Abstract] OR "Wells"[Title/Abstract] OR "Westminster"[Title/Abstract] OR "Winchester"[Title/Abstract] OR "Wolverhampton"[Title/Abstract] OR "Worcester"[Title/Abstract] OR "Wrexham"[Title/Abstract] OR "York"[Title/Abstract])) OR (UK[Affiliation])) OR (UK[Title/Abstract])) OR (England[Affiliation])) OR (England[Title/Abstract])) OR (Wales[Affiliation])) OR (Wales[Title/Abstract])) OR (Scotland[Title/Abstract])) OR (Scotland[Affiliation])) OR ("Northern Ireland"[Affiliation])) OR ("Northern Ireland"[Title/Abstract])) OR ("Britain"[Title/Abstract])) OR (Britain[Affiliation])) AND ("ethnicity"[Text Word] OR "ethnic group"[Text Word] OR "Black African"[Text Word] OR "Black Caribbean"[Text Word] OR "Mixed African"[Text Word] OR "Mixed Caribbean"[Text Word] OR "Afro-Caribbean"[Text Word] OR "black woman"[Text Word] OR "black women"[Text Word] OR "black race"[Text Word] OR "black ethnicity"[Text Word] OR "African heritage"[Text Word] OR "Caribbean heritage"[Text Word] OR "black heritage"[Text Word] OR "mixed black"[Text Word] OR "African descent"[Text Word] OR "Caribbean descent"[Text Word] OR "Afro-Caribbean descent"[Text Word] OR "African ancestry"[Text Word] OR "Caribbean ancestry"[Text Word] OR "Afro-Caribbean ancestry"[Text Word] OR "underserved"[All Fields] OR "ethnic minority"[Text Word] OR "ethnic minorities"[Text Word] OR "people of color"[Text Word] OR "people of colour"[Text Word] OR "global majority"[Text Word] OR "racial minority"[Text Word] OR "racial minorities"[Text Word])) AND ("Fibroids"[Text Word] OR "Myomas"[Text Word] OR "Leiomyomas"[Text Word] OR "uterine tumor"[Text Word] OR "uterine tumour"[Text Word] OR "Levonorgestrel intrauterine system"[Text Word] OR "LNG-IUS"[Text Word] OR "Tranexamic acid"[Text Word] OR "progestogen"[Text Word] OR "Gonadotropin releasing hormone analogues"[Text Word] OR "GnRHas"[Text Word] OR "Ulipristal acetate"[Text Word] OR "Hysterectomy"[Text Word] OR "Myomectomy"[Text Word] OR "Hysteroscopic resection"[Text Word] OR "Hysteroscopic morcellation"[Text Word] OR "Uterine artery embolisation"[Text Word] OR "Uterine artery embolization"[Text Word] OR "Endometrial ablation"[Text Word] OR "myometrium"[Text Word] OR "uterine myocytes"[Text Word] OR "Leiomyomata uteri"[Text Word] OR "fibromyomata uteri"[Text Word]) | 207 |
| 15 | "Fibroids"[Text Word] OR "Myomas"[Text Word] OR "Leiomyomas"[Text Word] OR "uterine tumor"[Text Word] OR "uterine tumour"[Text Word] OR "Levonorgestrel intrauterine system"[Text Word] OR "LNG-IUS"[Text Word] OR "Tranexamic acid"[Text Word] OR "progestogen"[Text Word] OR "Gonadotropin releasing hormone analogues"[Text Word] OR "GnRHas"[Text Word] OR "Ulipristal acetate"[Text Word] OR "Hysterectomy"[Text Word] OR "Myomectomy"[Text Word] OR "Hysteroscopic resection"[Text Word] OR "Hysteroscopic morcellation"[Text Word] OR "Uterine artery embolisation"[Text Word] OR "Uterine artery embolization"[Text Word] OR "Endometrial ablation"[Text Word] OR "myometrium"[Text Word] OR "uterine myocytes"[Text Word] OR "Leiomyomata uteri"[Text Word] OR "fibromyomata uteri"[Text Word] | 96,244 |
| 14 | "ethnicity"[Text Word] OR "ethnic group"[Text Word] OR "Black African"[Text Word] OR "Black Caribbean"[Text Word] OR "Mixed African"[Text Word] OR "Mixed Caribbean"[Text Word] OR "Afro-Caribbean"[Text Word] OR "black woman"[Text Word] OR "black women"[Text Word] OR "black race"[Text Word] OR "black ethnicity"[Text Word] OR "African heritage"[Text Word] OR "Caribbean heritage"[Text Word] OR "black heritage"[Text Word] OR "mixed black"[Text Word] OR "African descent"[Text Word] OR "Caribbean descent"[Text Word] OR "Afro-Caribbean descent"[Text Word] OR "African ancestry"[Text Word] OR "Caribbean ancestry"[Text Word] OR "Afro-Caribbean ancestry"[Text Word] OR "underserved"[All Fields] OR "ethnic minority"[Text Word] OR "ethnic minorities"[Text Word] OR "people of color"[Text Word] OR "people of colour"[Text Word] OR "global majority"[Text Word] OR "racial minority"[Text Word] OR "racial minorities"[Text Word] | 224,896 |
| 13 | ((((((((((((("Aberdeen"[Affiliation] OR "Armagh"[Affiliation] OR "Bangor"[Affiliation] OR "Bangor"[Affiliation] OR "Bath"[Affiliation] OR "Belfast"[Affiliation] OR "Birmingham"[Affiliation] OR "Bradford"[Affiliation] OR "Brighton & Hove"[Affiliation] OR "Bristol"[Affiliation] OR "Cambridge"[Affiliation] OR "Canterbury"[Affiliation] OR "Cardiff"[Affiliation] OR "Carlisle"[Affiliation] OR "Chelmsford"[Affiliation] OR "Chester"[Affiliation] OR "Chichester"[Affiliation] OR "Colchester"[Affiliation] OR "Coventry"[Affiliation] OR "Derby"[Affiliation] OR "Doncaster"[Affiliation] OR "Dundee"[Affiliation] OR "Dunfermline"[Affiliation] OR "Durham"[Affiliation] OR "Edinburgh"[Affiliation] OR "Ely"[Affiliation] OR "England"[Affiliation] OR "Exeter"[Affiliation] OR "Glasgow"[Affiliation] OR "Gloucester"[Affiliation] OR "Hereford"[Affiliation] OR "Inverness"[Affiliation] OR "Kingston-Upon-Hull"[Affiliation] OR "Lancaster"[Affiliation] OR "Leeds"[Affiliation] OR "Leicester"[Affiliation] OR "Lichfield"[Affiliation] OR "Lincoln"[Affiliation] OR "Lisburn"[Affiliation] OR "Liverpool"[Affiliation] OR "London"[Affiliation] OR "Londonderry"[Affiliation] OR "Manchester"[Affiliation] OR "Milton Keynes"[Affiliation] OR "Newcastle-Upon-Tyne"[Affiliation] OR "Newport"[Affiliation] OR "Newry"[Affiliation] OR "Northern Ireland"[Affiliation] OR "Norwich"[Affiliation] OR "Nottingham"[Affiliation] OR "Oxford"[Affiliation] OR "Perth"[Affiliation] OR "Peterborough"[Affiliation] OR "Plymouth"[Affiliation] OR "Portsmouth"[Affiliation] OR "Preston"[Affiliation] OR "Ripon"[Affiliation] OR "Salford"[Affiliation] OR "Salisbury"[Affiliation] OR "Scotland"[Affiliation] OR "Sheffield"[Affiliation] OR "Southampton"[Affiliation] OR "Southend-on-Sea"[Affiliation] OR "St Albans"[Affiliation] OR "St Asaph"[Affiliation] OR "St Davids"[Affiliation] OR "Stirling"[Affiliation] OR "Stoke on Trent"[Affiliation] OR "Sunderland"[Affiliation] OR "Swansea"[Affiliation] OR "Truro"[Affiliation] OR "United Kingdom"[Affiliation] OR "Wakefield"[Affiliation] OR "Wales"[Affiliation] OR "Wells"[Affiliation] OR "Westminster"[Affiliation] OR "Winchester"[Affiliation] OR "Wolverhampton"[Affiliation] OR "Worcester"[Affiliation] OR "Wrexham"[Affiliation] OR "York"[Affiliation]) OR ("Aberdeen"[Title/Abstract] OR "Armagh"[Title/Abstract] OR "Bangor"[Title/Abstract] OR "Bath"[Title/Abstract] OR "Belfast"[Title/Abstract] OR "Birmingham"[Title/Abstract] OR "Bradford"[Title/Abstract] OR "Brighton hove"[Title/Abstract] OR "Bristol"[Title/Abstract] OR "Cambridge"[Title/Abstract] OR "Canterbury"[Title/Abstract] OR "Cardiff"[Title/Abstract] OR "Carlisle"[Title/Abstract] OR "Chelmsford"[Title/Abstract] OR "Chester"[Title/Abstract] OR "Chichester"[Title/Abstract] OR "Colchester"[Title/Abstract] OR "Coventry"[Title/Abstract] OR "Derby"[Title/Abstract] OR "Doncaster"[Title/Abstract] OR "Dundee"[Title/Abstract] OR "Dunfermline"[Title/Abstract] OR "Durham"[Title/Abstract] OR "Edinburgh"[Title/Abstract] OR "Ely"[Title/Abstract] OR "England"[Title/Abstract] OR "Exeter"[Title/Abstract] OR "Glasgow"[Title/Abstract] OR "Gloucester"[Title/Abstract] OR "Hereford"[Title/Abstract] OR "Inverness"[Title/Abstract] OR "Kingston-Upon-Hull"[Title/Abstract] OR "Lancaster"[Title/Abstract] OR "Leeds"[Title/Abstract] OR "Leicester"[Title/Abstract] OR "Lichfield"[Title/Abstract] OR "Lincoln"[Title/Abstract] OR "Lisburn"[Title/Abstract] OR "Liverpool"[Title/Abstract] OR "London"[Title/Abstract] OR "Londonderry"[Title/Abstract] OR "Manchester"[Title/Abstract] OR "Milton Keynes"[Title/Abstract] OR "Newcastle-Upon-Tyne"[Title/Abstract] OR "Newport"[Title/Abstract] OR "Newry"[Title/Abstract] OR "Northern Ireland"[Title/Abstract] OR "Norwich"[Title/Abstract] OR "Nottingham"[Title/Abstract] OR "Oxford"[Title/Abstract] OR "Perth"[Title/Abstract] OR "Peterborough"[Title/Abstract] OR "Plymouth"[Title/Abstract] OR "Portsmouth"[Title/Abstract] OR "Preston"[Title/Abstract] OR "Ripon"[Title/Abstract] OR "Salford"[Title/Abstract] OR "Salisbury"[Title/Abstract] OR "Scotland"[Title/Abstract] OR "Sheffield"[Title/Abstract] OR "Southampton"[Title/Abstract] OR "Southend-on-Sea"[Title/Abstract] OR "St Albans"[Title/Abstract] OR "St Davids"[Title/Abstract] OR "Stirling"[Title/Abstract] OR "Stoke on Trent"[Title/Abstract] OR "Sunderland"[Title/Abstract] OR "Swansea"[Title/Abstract] OR "Truro"[Title/Abstract] OR "United Kingdom"[Title/Abstract] OR "Wakefield"[Title/Abstract] OR "Wales"[Title/Abstract] OR "Wells"[Title/Abstract] OR "Westminster"[Title/Abstract] OR "Winchester"[Title/Abstract] OR "Wolverhampton"[Title/Abstract] OR "Worcester"[Title/Abstract] OR "Wrexham"[Title/Abstract] OR "York"[Title/Abstract])) OR (UK[Affiliation])) OR (UK[Title/Abstract])) OR (England[Affiliation])) OR (England[Title/Abstract])) OR (Wales[Affiliation])) OR (Wales[Title/Abstract])) OR (Scotland[Title/Abstract])) OR (Scotland[Affiliation])) OR ("Northern Ireland"[Affiliation])) OR ("Northern Ireland"[Title/Abstract])) OR ("Britain"[Title/Abstract])) OR (Britain[Affiliation]) | 3,770,501 |
| 12 | Britain[Affiliation] | 5,536 |
| 11 | "Britain"[Title/Abstract] | 20,620 |
| 10 | "Northern Ireland"[Title/Abstract] | 6,254 |
| 9 | "Northern Ireland"[Affiliation] | 18,846 |
| 8 | Scotland[Affiliation] | 59,642 |
| 7 | Scotland[Title/Abstract] | 20,534 |
| 6 | Wales[Title/Abstract] | 30,037 |
| 5 | Wales[Affiliation] | 178,865 |
| 4 | England[Title/Abstract] | 68,180 |
| 3 | England[Affiliation] | 104,999 |
| 2 | UK[Title/Abstract] | 161,063 |
| 1 | UK[Affiliation] | 1,502,353 |

**CINAHL (n=76)**

| **S4** | S1 AND S2 AND S3 | 76 |
| --- | --- | --- |
| **S3** | TX ( "Aberdeen" OR "Armagh" OR "Bangor" OR "Bath" OR "Belfast" OR "Birmingham" OR "Bradford" OR "Brighton hove" OR "Bristol" OR "Cambridge" OR "Canterbury" OR "Cardiff" OR "Carlisle" OR "Chelmsford" OR "Chester" OR "Chichester" OR "Colchester" OR "Coventry" OR "Derby" OR "Doncaster" OR "Dundee" OR "Dunfermline" OR "Durham" OR "Edinburgh" OR "Ely" OR "England" OR "Exeter" OR "Glasgow" OR "Gloucester" OR "Hereford" OR "Inverness" OR "Kingston-Upon-Hull" OR "Lancaster" OR "Leeds" OR "Leicester" OR "Lichfield" OR "Lincoln" OR "Lisburn" OR "Liverpool" OR "London" OR "Londonderry" OR "Manchester" OR "Milton Keynes" OR "Newcastle-Upon-Tyne" OR "Newport" OR "Newry" OR "Northern Ireland" OR "Norwich" OR "Nottingham" OR "Oxford" OR "Perth" OR "Peterborough" OR "Plymouth" OR "Portsmouth" OR "Preston" OR "Ripon" OR "Salford" OR "Salisbury" OR "Scotland" OR "Sheffield" OR "Southampton" OR "Southend-on-Sea" OR "St Albans" OR "St Asaph" OR "St Davids" OR "Stirling" OR "Stoke on Trent" OR "Sunderland" OR "Swansea" OR "Truro" OR "United Kingdom" OR "Wakefield" OR "Wales" OR "Wells" OR "Westminster" OR "Winchester" OR "Wolverhampton" OR "Worcester" OR "Wrexham" OR "York" OR "Aberdeen" OR "Armagh" OR "Bangor" OR "Bath" OR "Belfast" OR "Birmingham" OR "Bradford" OR "Brighton hove" OR "Bristol" OR "Cambridge" OR "Canterbury" OR "Cardiff" OR "Carlisle" OR "Chelmsford" OR "Chester" OR "Chichester" OR "Colchester" OR "Coventry" OR "Derby" OR "Doncaster" OR "Dundee" OR "Dunfermline" OR "Durham" OR "Edinburgh" OR "Ely" OR "England" OR "Exeter" OR "Glasgow" OR "Gloucester" OR "Hereford" OR "Inverness" OR "Kingston-Upon-Hull" OR "Lancaster" OR "Leeds" OR "Leicester" OR "Lichfield" OR "Lincoln" OR "Lisburn" OR "Liverpool" OR "London" OR "Londonderry" OR "Manchester" OR "Milton Keynes" OR "Newcastle-Upon-Tyne" OR "Newport" OR "Newry" OR "Northern Ireland" OR "Norwich" OR "Nottingham" OR "Oxford" OR "Perth" OR "Peterborough" OR "Plymouth" OR "Portsmouth" OR "Preston" OR "Ripon" OR "Salford" OR "Salisbury" OR "Scotland" OR "Sheffield" OR "Southampton" OR "Southend-on-Sea" OR "St Albans" OR "St Davids" OR "Stirling" OR "Stoke on Trent" OR "Sunderland" OR "Swansea" OR "Truro" OR "United Kingdom" OR "Wakefield" OR "Wales" OR "Wells" OR "Westminster" OR "Winchester" OR "Wolverhampton" OR "Worcester" OR "Wrexham" OR "York" OR "UK" OR "UK" OR "England" OR "England" OR "Wales" OR "Wales" OR "Scotland" OR "Scotland" OR "Northern Ireland" OR "Northern Ireland" OR "Britain" OR "Britain" OR "ICS" OR "ICB" OR "GP" OR "Hospital" OR "clinic" OR "integrated care board" OR "integrated care system" OR "NHS" ) OR XB ( "Aberdeen" OR "Armagh" OR "Bangor" OR "Bath" OR "Belfast" OR "Birmingham" OR "Bradford" OR "Brighton hove" OR "Bristol" OR "Cambridge" OR "Canterbury" OR "Cardiff" OR "Carlisle" OR "Chelmsford" OR "Chester" OR "Chichester" OR "Colchester" OR "Coventry" OR "Derby" OR "Doncaster" OR "Dundee" OR "Dunfermline" OR "Durham" OR "Edinburgh" OR "Ely" OR "England" OR "Exeter" OR "Glasgow" OR "Gloucester" OR "Hereford" OR "Inverness" OR "Kingston-Upon-Hull" OR "Lancaster" OR "Leeds" OR "Leicester" OR "Lichfield" OR "Lincoln" OR "Lisburn" OR "Liverpool" OR "London" OR "Londonderry" OR "Manchester" OR "Milton Keynes" OR "Newcastle-Upon-Tyne" OR "Newport" OR "Newry" OR "Northern Ireland" OR "Norwich" OR "Nottingham" OR "Oxford" OR "Perth" OR "Peterborough" OR "Plymouth" OR "Portsmouth" OR "Preston" OR "Ripon" OR "Salford" OR "Salisbury" OR "Scotland" OR "Sheffield" OR "Southampton" OR "Southend-on-Sea" OR "St Albans" OR "St Asaph" OR "St Davids" OR "Stirling" OR "Stoke on Trent" OR "Sunderland" OR "Swansea" OR "Truro" OR "United Kingdom" OR "Wakefield" OR "Wales" OR "Wells" OR "Westminster" OR "Winchester" OR "Wolverhampton" OR "Worcester" OR "Wrexham" OR "York" OR "Aberdeen" OR "Armagh" OR "Bangor" OR "Bath" OR "Belfast" OR "Birmingham" OR "Bradford" OR "Brighton hove" OR "Bristol" OR "Cambridge" OR "Canterbury" OR "Cardiff" OR "Carlisle" OR "Chelmsford" OR "Chester" OR "Chichester" OR "Colchester" OR "Coventry" OR "Derby" OR "Doncaster" OR "Dundee" OR "Dunfermline" OR "Durham" OR "Edinburgh" OR "Ely" OR "England" OR "Exeter" OR "Glasgow" OR "Gloucester" OR "Hereford" OR "Inverness" OR "Kingston-Upon-Hull" OR "Lancaster" OR "Leeds" OR "Leicester" OR "Lichfield" OR "Lincoln" OR "Lisburn" OR "Liverpool" OR "London" OR "Londonderry" OR "Manchester" OR "Milton Keynes" OR "Newcastle-Upon-Tyne" OR "Newport" OR "Newry" OR "Northern Ireland" OR "Norwich" OR "Nottingham" OR "Oxford" OR "Perth" OR "Peterborough" OR "Plymouth" OR "Portsmouth" OR "Preston" OR "Ripon" OR "Salford" OR "Salisbury" OR "Scotland" OR "Sheffield" OR "Southampton" OR "Southend-on-Sea" OR "St Albans" OR "St Davids" OR "Stirling" OR "Stoke on Trent" OR "Sunderland" OR "Swansea" OR "Truro" OR "United Kingdom" OR "Wakefield" OR "Wales" OR "Wells" OR "Westminster" OR "Winchester" OR "Wolverhampton" OR "Worcester" OR "Wrexham" OR "York" OR "UK" OR "UK" OR "England" OR "England" OR "Wales" OR "Wales" OR "Scotland" OR "Scotland" OR "Northern Ireland" OR "Northern Ireland" OR "Britain" OR "Britain" OR "ICS" OR "ICB" OR "GP" OR "Hospital" OR "clinic" OR "integrated care board" OR "integrated care system" OR "NHS" ) | 5,511,069 |
| **S2** | TX ( "Fibroids" OR "Myomas" OR "Leiomyomas" OR "uterine tumor" OR "uterine tumour" OR "Levonorgestrel intrauterine system" OR "LNG-IUS" OR "Tranexamic acid" OR "progestogen" OR "Gonadotropin releasing hormone analogues" OR "GnRHas" OR "Ulipristal acetate" OR "Hysterectomy" OR "Myomectomy" OR "Hysteroscopic resection" OR "Hysteroscopic morcellation" OR "Uterine artery embolisation" OR "Uterine artery embolization" OR "Endometrial ablation" OR "myometrium" OR "uterine myocytes" OR "Leiomyomata uteri" OR "fibromyomata uteri" ) OR XB ( "Fibroids" OR "Myomas" OR "Leiomyomas" OR "uterine tumor" OR "uterine tumour" OR "Levonorgestrel intrauterine system" OR "LNG-IUS" OR "Tranexamic acid" OR "progestogen" OR "Gonadotropin releasing hormone analogues" OR "GnRHas" OR "Ulipristal acetate" OR "Hysterectomy" OR "Myomectomy" OR "Hysteroscopic resection" OR "Hysteroscopic morcellation" OR "Uterine artery embolisation" OR "Uterine artery embolization" OR "Endometrial ablation" OR "myometrium" OR "uterine myocytes" OR "Leiomyomata uteri" OR "fibromyomata uteri" ) | 25,178 |
| **S1** | TX ( "ethnicity" OR "ethnic group" OR "Black African" OR "Black Caribbean" OR "Mixed African" OR "Mixed Caribbean" OR "Afro-caribbean" OR "black woman" OR "black women" OR "black race" OR "black ethnicity" OR "african heritage" OR "caribbean heritage" OR "black heritage" OR "mixed black" OR "african descent" OR "caribbean descent" OR "afro-caribbean descent" OR "african ancestry" OR "caribbean ancestry" OR "afro-caribbean ancestry" OR "underserved" OR "ethnic minority" OR "ethnic minorities" OR "people of color" OR "people of colour" OR "global majority" OR "racial minority" OR "racial minorities" ) OR XB ( "ethnicity" OR "ethnic group" OR "Black African" OR "Black Caribbean" OR "Mixed African" OR "Mixed Caribbean" OR "Afro-caribbean" OR "black woman" OR "black women" OR "black race" OR "black ethnicity" OR "african heritage" OR "caribbean heritage" OR "black heritage" OR "mixed black" OR "african descent" OR "caribbean descent" OR "afro-caribbean descent" OR "african ancestry" OR "caribbean ancestry" OR "afro-caribbean ancestry" OR "underserved" OR "ethnic minority" OR "ethnic minorities" OR "people of color" OR "people of colour" OR "global majority" OR "racial minority" OR "racial minorities" ) | 86,927 |

**Cochrane (n=3)**

| **4** | #1 AND #2 AND #3 | 3 |
| --- | --- | --- |
| **3** | ("Fibroids"):ti,ab,kw OR ("Myomas"):ti,ab,kw OR ("Leiomyomas"):ti,ab,kw OR ("uterine tumor"):ti,ab,kw OR ("uterine tumour"):ti,ab,kw OR ("Levonorgestrel intrauterine system"):ti,ab,kw OR ("LNG-IUS"):ti,ab,kw OR ("Tranexamic acid"):ti,ab,kw OR ("progestogen"):ti,ab,kw OR ("Gonadotropin releasing hormone analogues"):ti,ab,kw OR ("GnRHas"):ti,ab,kw OR ("Ulipristal acetate"):ti,ab,kw OR ("Hysterectomy"):ti,ab,kw OR ("Myomectomy"):ti,ab,kw OR ("Hysteroscopic resection"):ti,ab,kw OR ("Hysteroscopic morcellation"):ti,ab,kw OR ("Uterine artery embolisation"):ti,ab,kw OR ("Uterine artery embolization"):ti,ab,kw OR ("Endometrial ablation"):ti,ab,kw OR ("myometrium"):ti,ab,kw OR ("uterine myocytes"):ti,ab,kw | 17534 |
| **2** | ("England"):ti,ab,kw OR ("United Kingdom"):ti,ab,kw OR ("Scotland"):ti,ab,kw OR ("Wales"):ti,ab,kw OR ("Northern Ireland"):ti,ab,kw OR ("England"):ti,ab,kw OR ("United kingdom"):ti,ab,kw OR ("Scotland"):ti,ab,kw OR ("Wales"):ti,ab,kw OR ("Northern Ireland"):ti,ab,kw OR ("Britain"):ti,ab,kw OR ("London"):ti,ab,kw OR ("Manchester"):ti,ab,kw OR ("Cardiff"):ti,ab,kw OR ("Edinburgh"):ti,ab,kw OR ("ICS"):ti,ab,kw OR ("ICB"):ti,ab,kw OR ("GP"):ti,ab,kw OR ("Hospital"):ti,ab,kw OR ("clinic"):ti,ab,kw OR ("integrated care board"):ti,ab,kw OR ("integrated care system"):ti,ab,kw | 272269 |
| **1** | ("ethnicity"):ti,ab,kw OR ("ethnic group"):ti,ab,kw OR ("Black African"):ti,ab,kw OR ("Black Caribbean"):ti,ab,kw OR ("Mixed African"):ti,ab,kw OR ("Mixed Caribbean"):ti,ab,kw OR ("Afro-caribbean"):ti,ab,kw OR ("black woman"):ti,ab,kw OR ("black women"):ti,ab,kw OR ("black race"):ti,ab,kw OR ("black ethnicity"):ti,ab,kw OR ("african heritage"):ti,ab,kw OR ("caribbean heritage"):ti,ab,kw OR ("black heritage"):ti,ab,kw OR ("mixed black"):ti,ab,kw OR ("african descent"):ti,ab,kw OR ("caribbean descent"):ti,ab,kw OR ("afro-caribbean descent"):ti,ab,kw OR ("african ancestry"):ti,ab,kw OR ("caribbean ancestry"):ti,ab,kw OR ("afro-caribbean ancestry"):ti,ab,kw OR ("underserved"):ti,ab,kw OR ("ethnic minority"):ti,ab,kw OR ("ethnic minorities"):ti,ab,kw OR ("people of color"):ti,ab,kw OR ("people of colour"):ti,ab,kw OR ("global majority"):ti,ab,kw OR (“racial minority”):ti,ab,kw OR (“racial minorities”):ti,ab,kw | 14798 |
